# Pathways of Exposure to *Vibrio Cholerae* in an Urban Informal Settlement in Nairobi, Kenya

**DOI:** 10.1101/2024.01.17.24301425

**Authors:** Kelvin Kering, Yuke Wang, Cecilia Mbae, Michael Mugo, Beatrice Ongadi, Georgina Odityo, Peter Muturi, Habib Yakubu, Pengbo Liu, Sarah Durry, Aniruddha Deshpande, Wondwossen Gebreyes, Christine Moe, Samuel Kariuki

**Affiliations:** Centre for Microbiology Research, Kenya Medical Research Institute, Nairobi, Kenya; Center for Global Safe Water, Sanitation, and Hygiene, Hubert Department of Global Health, Rollins School of Public Health, Emory University, Atlanta, GA, USA; Department of Epidemiology, Rollins School of Public Health, Emory University, Atlanta, Georgia; Global One Health initiative (GOHi), The Ohio State University, Columbus, OH, USA; Veterinary Preventive Medicine, The Ohio State University, Columbus, OH, USA

**Keywords:** *Vibrio cholerae*, Environmental samples, Exposure assessment, Informal settlement, Kenya

## Abstract

Cholera is a diarrhoeal disease caused by the toxigenic *Vibrio cholerae* (*V. cholerae*) bacterium. *V. cholerae* can contaminate drinking water sources and food through poor sanitation and hygiene, especially in informal settlements and refugee camps where cholera outbreaks have been reported in Kenya.

This study aimed to identify environmental transmission routes of *V. cholerae* within Mukuru informal settlement in Nairobi. We collected nine types of environmental samples (drinking water, flood water, open drains, surface water, shaved ice, raw produce, street food, soil, and public latrine swabs) over 12 months. All samples were analysed for *V. cholerae* by culture and qPCR, then qPCR-positive samples were quantified using a *V. cholerae* DNA standard. Behavioural data was collected to determine the frequency of contact with the environment among adults and children.

Of the 803 samples collected, 20.4% were positive for *V. cholerae* by qPCR. However, none were positive for *V. cholerae* by culture. *V. cholerae* genes were detected in the majority of the environmental water samples (79.3%), including open drains, flood water, and surface water, but were only detected in small proportions of other sample types. Vibrio-positive environmental water samples had higher mean *V. cholerae* concentrations [2490–3469 genome copies (gc) per millilitre (mL)] compared to drinking water samples (25.6 gc/mL). Combined with the behavioural data, exposure assessment showed that contact with surface water had the highest contribution to the total *V. cholerae* exposure among children while ingestion of municipal drinking water and street food and contact with surface water made substantial contributions to the total *V. cholerae* exposure for adults.

Detection of *V. cholerae* in street food and drinking water indicates risk of both endemic and epidemic cholera. Exposure to *V. cholerae* through multiple pathways highlights the need to improve water and sanitation infrastructure, strengthen food hygiene practices, and roll out cholera vaccination.

## Introduction

Cholera is an acute infectious diarrhoeal disease caused by toxigenic *Vibrio cholerae* (*V. cholerae*), a comma-shaped, gram-negative bacterial pathogen [1,2] transmitted to humans through the consumption of contaminated food and water. It is estimated that approximately 2.9 million cases of cholera occur annually, resulting in 95,000 deaths globally, with low- and middle-income countries bearing the biggest burden [3]. Of the cholera cases reported to the World Health Organization (WHO) between 1996 and 2019 [4,5] the majority were reported from sub-Saharan Africa. In Kenya, cholera was first reported in 1971, with several outbreaks being reported every five to seven years [6,7]. Cholera cases are often recorded in refugee camps [8,9] and Nairobi’s urban informal settlements [10] with case fatality rates (CFR) greater than 2.5%. For instance, in 2016, cholera cases were reported in 22 of Kenya’s 47 counties [9] with an average CFR of 1.6 [7], while in 2019, 5208 cases were recorded, with a CFR of 2.7% [7]. The high CFR observed in Kenya is concerning and highlights the need to map cholera hotspots, identify risk factors, and implement evidence-based integrated intervention strategies.

*V. cholerae* is pervasive in aquatic environments and thrives as free-living cells in brackish and freshwater or biofilms in zooplankton and phytoplankton [2,11]. However, in unfavourable environmental conditions and limited nutrition, *V. cholerae* enter a viable but non-culturable (VBNC) state [12–14]. From aquatic environments and environmental reservoirs, *V. cholerae* can spread into drinking water sources and food through contaminated runoff from flooding and/or wastewater [15], thereby exposing the human population. The presence and persistence of *V. cholerae* in environmental reservoirs (freshwater, wastewater, rivers) [16–18] enables microbial risk assessment using data on human behaviour interactions with those reservoirs and the levels of *V. cholerae* in these environments.

Impoverished populations, especially in informal settlements [6], refugee camps, and areas afflicted by natural calamities and conflicts [19] are disproportionately affected by cholera due to poor water, sanitation, and hygiene (WASH), limited access to healthcare, and weak healthcare systems. Due to the rapid population growth and unplanned residential developments, it is estimated that as of 2023 about 60% of Kenya’s population resides in informal settlements and slums [20,21], some of which are located within Kenya’s capital, Nairobi. One of the informal settlements in Nairobi, Mukuru Slums, is characterized by overcrowding, poor solid-waste management, inadequate WASH infrastructure and limited access to healthcare. While there have been previous reports of cholera outbreaks within the Mukuru informal settlements, the probable pathways of exposure have not been identified. SaniPath is a multi-pathway exposure assessment approach that identifies and characterizes the pathways of exposure to fecal contamination and enables the prioritization and design of WASH interventions [22,23]. This study extended the SaniPath approach for assessing exposure to fecal contamination to exposure specifically to *V. cholerae.* Our goal was to understand the transmission of *V. cholerae* by examining plausible pathways of exposure to *V. cholerae* and examining spatial and temporal variation in *V. cholerae* detection and concentration in different environmental reservoirs. The results from this study will guide the development of evidence-based integrated prevention and control strategies for this cholera-endemic setting.

## Materials and Methods

### Study site

The study was conducted in the Mukuru informal settlement, which is located 20 km east of Nairobi in Kenya. This informal settlement is divided into eight villages and this study was conducted in two villages (Mukuru Kwa Njenga and Mukuru Kwa Reuben), hereafter referred to as neighbourhoods. The neighbourhoods are further divided into zones. Based on local hospital records (from the Mukuru Kwa Reuben Health Centre, Mukuru Health Centre, and Medical Missionaries of Mary) and community health volunteers (CHVs) reports of previous cholera cases, the study was conducted in five zones identified as cholera hotspots (zones that had reported cholera cases in the most recent outbreak, 2018) [24]. Three zones (Gatoto, Mombasa, Feed the Children-FTC) within Mukuru kwa Reuben and two zones (Wapewape and Pipeline) within Mukuru kwa Njenga neighbourhood were selected as study sites (**Figure 1**).

**Figure 1:**
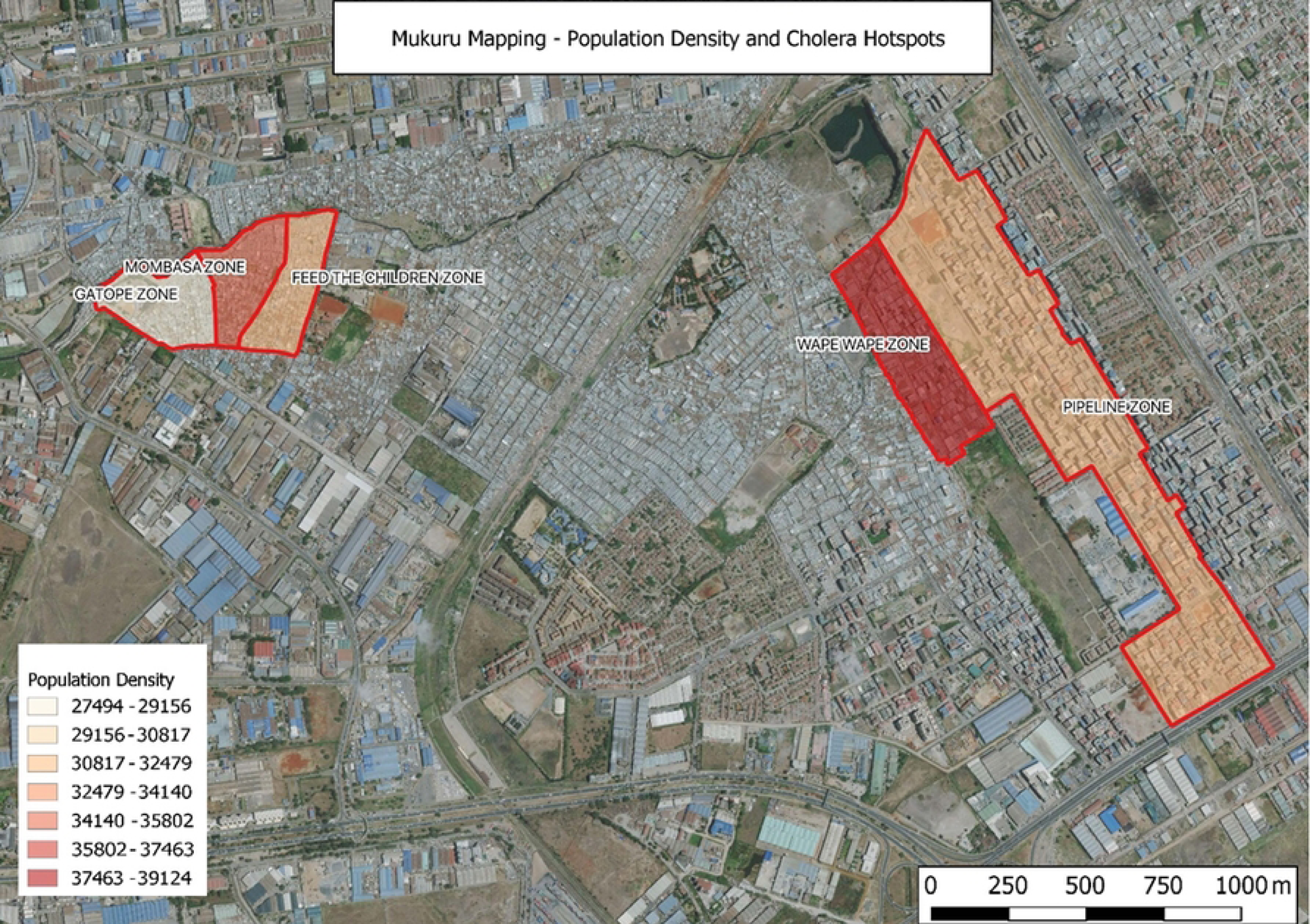
Mapped cholera hotspots (Gatope, Mombasa, and Feed the Children) in the Mukuru Kwa Reuben neighbourhood and (Wapewape and Pipeline) in the Mukuru Kwa Njenga neighbourhood. The unit for population density is persons per square kilometre.

### Identification of transmission routes

Transect walks and information from CHVs were used to identify plausible exposure pathways and transmission routes for *V. cholerae*. The transect walks were conducted as previously described [25,26]. The information provided by the CHVs included the pathways that are relevant to the Mukuru informal settlement and the most appropriate sampling sites. The sampling sites were selected in areas where the residents often interacted with those pathways, including the children’s playgrounds, the most frequently used public sanitation facilities, common public water sources, and the most commonly consumed produce from markets and street foods from neighbourhood street vendors.

### Behaviour survey and environmental sample collection

Between November 2020 and April 2022, we conducted 400 household surveys from randomly selected households, 16 community surveys (average of 18 participants per survey), and 16 school surveys (20 participants per survey) in the study neighbourhoods using the SaniPath Tool [25,27]. Respondents to the household surveys were adults residing within the two study neighbourhoods who were involved in the management of water within the household and familiar with children’s activities. The community and school surveys were conducted separately for male and female participants. Participants in the community survey were persons residing within the two neighbourhoods and with a child aged 5–12 years. The school surveys were conducted in four schools in the two neighbourhoods with school children aged 10– 12 years. The surveys collected behaviour information related to the frequency of contact with different compartments of the environment (eg. open drains, flood water, surface water, drinking water source, consumption of street food and raw produce, etc.) by adults and children.

In the two neighbourhoods, ten sampling sites (five per neighbourhood) were randomly selected for each sample type. Six sample types (public latrine swabs, soil, surface water, flood water, raw produce, and shaved ice) were collected weekly for 4 months (July– November 2021), the other three sample types (drinking water, open drain, and street food) were collected weekly for one year (July 2021–July 2022). Samples were collected as previously described [26]. Briefly, 1) soil was collected from common children’s playgrounds and public gathering areas; 2) surface water from areas along the rivers where there was frequent human activity or river crossing; 3) public latrines wall surfaces (100 cm^2^) that are frequently touched and door handles were swabbed using two swabs per latrine; 4) flood water was collected from stagnant water in areas with human activity; 5) open drain water was collected from concrete and unlined open drainage channels containing liquid and solid waste, rain, flood water, and wastewater from toilets and household activities in areas where reported human contact was frequent; 6) drinking water was collected from communal municipal water points, boreholes, water vendors, and stored drinking water from randomly selected households where household surveys were conducted; 7) street food reported to be commonly consumed by neighbourhood children and adults, 8) produce reported to be commonly consumed raw, and 9) shaved ice purchased from vendors.

Prior to sampling drinking water, chlorine concentration was determined. Drinking water containing chlorine was collected directly from the source into 100 mL Whirl-pak bags containing sodium thiosulphate. Surface water, flood water, and open drain water samples were collected using a sterile metal bucket. The bucket was then placed on the ground to allow the water to settle for five minutes, thereafter the water was poured into 2L Whirl-pak bags. Soil samples were collected at a 45° angle to a depth of 5cm using a sterile spatula. Hard-packed soil samples were gently broken using the spatula before placing them in 100 mL Whirl-pak bags. Street food was collected from vendors according to the quantity normally served to an individual. Raw produce samples included tomato and coriander, and were collected and placed into 2L Whirl-pak bags. The shaved ice was also collected in quantities typically consumed by an individual and placed in Whirl-pak bags. Public latrine surfaces were swabbed using Puritan environmental sampling swabs pre-moistened with neutralizing buffer and 0.1% peptone water (EnviroMax Plus, Puritan Medical Products Company LLC, Maine, USA). The Whirl-pak bags containing the samples were then placed in cooler boxes containing ice packs and transported to the Microbiology Laboratory at KEMRI and processed within six hours of sample collection.

### Microbiological culture of samples for *Vibrio cholerae*

All samples were initially enriched in Alkaline Peptone Water (APW). For surface water, flood water, open drain water, drinking water, and shaved ice, 5 mL of sample was mixed with 5 mL of APW in 50 mL conical tubes and incubated at 37°C for 18 hours. For raw produce, street food, and latrine swabs, samples were rinsed and then 5 mL of the rinse solution was mixed with 20 mL of APW and incubated. Soil was initially mixed with distilled water and then 5 mL of the soil suspension was mixed with APW and incubated. Shaved ice was allowed to melt at room temperature before it was mixed with APW. The samples were then subcultured onto Thiosulfate Citrate Bile salts Sucrose (TCBS) agar and incubated at 37° C for 18 hours. Suspected *V. cholerae* colonies were subjected to both an oxidase test (Thermo Scientific Remel BactiDrop Oxidase) and *V. cholerae* 01 polyvalent antisera.

### Sample concentration

The samples were concentrated before DNA extraction. For drinking water (borehole water, water from vendors, and stored drinking water), 100 mL of each sample was filtered through a 0.45 µM membrane filter (Millipore). The membrane filters were then used as the template for DNA extraction. One hundred millilitres of open drain, flood water, or surface water was concentrated using 1% Bovine Serum Albumin (BSA), 12% Polyethylene Glycol (PEG), and 0.9M Sodium Chloride (NaCl). The street foods were initially rinsed with 100 mL of sterile water, after which the homogenized sample was concentrated using 1% BSA, 12% PEG, and 0.9M NaCl as described above. The raw produce was mixed with 100 mL of phosphate-buffered saline Tween (PBST), thereafter the solution was concentrated as described above. For soil, 100 g was mixed with 200 mL of distilled water by shaking vigorously for 30 minutes, and then sample was allowed to settle at room temperature for two hours. One hundred millilitres of the sample were then concentrated as described previously. Each public latrine swab was mixed with 7 mL of PBST, vortexed briefly, and allowed to incubate at room temperature for 5 minutes and then eluted with PBST. This was done twice for each of the two swabs collected from each public latrine. Approximately 15 mL of the recovered eluate was concentrated using BSA, PEG, and NaCl as described above. The concentrated samples and membrane filters were then stored at 4°C until further processing.

### DNA extraction

The samples were then centrifuged at 4200 revolutions per minute for one hour and the pellet was used for DNA extraction using the QIAamp FAST DNA stool Mini Kit (50) (Qiagen, Hilden, Germany), according to the manufacturer’s instructions. A known *Vibrio cholerae* isolate was used as a positive control, and sterile distilled water was used as the negative control. The extracted DNA was stored at −80 °C for further downstream analysis.

### Quantitative polymerase chain reaction (q-PCR) detection of *V. cholerae*

The presence of *V. cholerae* was examined through the detection of the hemolysin A (*hlyA*) gene by qPCR as described by Huang *et al* [28]. The 25 µL master-mix contained 12.5 µL Bio-Rad iQ Multiplex Powermix, 0.4 µM of each hlyA forward and reverse primers, 0.2 µM hlyA probe, 5 µL molecular water and 5 µL of DNA template. AMPLIRUN *Vibrio cholerae* DNA control (VIRCELL Microbiologists, MBC118) was used as the positive control and nuclease-free water was used as the negative control. Quantitative PCR was run on a Magnetic Induction Cycler (MIC) using the following conditions: activation (at 95 °C for 15 minutes) and 40 cycles of 95°C for 15 s, 55°C for 40 s and 72 °C for 30 s. PCR reactions were run in duplicate wells for each sample. Samples with both Ct values of ≤ 39 and a difference between the duplicate Ct values of < 2 were considered positive.

### Quantification of *V. cholerae* in positive samples

*V. cholerae* positive samples with two Ct values of ≤ 36 were subjected to quantitative detection of genome copies as previously described [29]. Briefly, commercially purchased AMPLIRUN *Vibrio cholerae* DNA standard (VIRCELL Microbiologists, MBC118) was serially diluted and subjected to qPCR to derive a standard curve. The standard curve was then used to quantify the genome copies in the positive samples. Quantification of genome copies in each sample was estimated by interpolation of the mean Ct value to the standard curve and the dilution factor for each sample type. Absolute equivalent genome copies was expressed as per mL for the water samples, per gram for the raw produce, soil, and street food samples, and per swab for the public latrine samples [29].

### Statistical Analysis

We summarized the percent of environmental samples with detected *V. cholerae* (positivity) and the mean concentration of *V. cholerae* among positive samples by pathway and neighbourhood. Temporal trends of the percent positive samples for each sample type, especially for drinking water, street food, and open drains, were examined. We conducted the SaniPath multi-pathway exposure assessment [25] for *V. cholerae* for adults and children and estimated exposure using the frequency of behaviour leading to contact with the environment and the concentration of *V. cholerae* in different environmental sample types. All the analyses were conducted in R version 4.0.1.

### Ethical Considerations

Ethical approval to conduct this study was sought and obtained from the Kenya Medical Research Institute review board; Scientific and Ethics Review Unit (**KEMRI/SERU/CMR/P00116/3871**), the National Commission for Science, Technology and Innovation (**NACOSTI/P/20/4566**) and the Nairobi City County (**CMO/NRB/OPR/VOL1-2/2020/32**). All adult participants were requested to participate in the surveys after providing written informed consent. In the case of school children, written informed consent was sought from their parents/guardians and the school administration.

## Results

### *V. cholerae* detection by environmental pathway

Based on the transect walks, nine sample types (**Table 1**) were identified as the most probable to be associated with exposure to *V. cholerae* within the Mukuru informal settlement. Few water vendors were found in Mukuru Kwa Reuben, therefore, a lesser number of water samples was collected from this location. Of the 803 environmental samples analyzed, none were culture-positive for *V. cholera*. However, 164 (20.4%) samples were positive for the *V. cholerae* hemolysin A gene by qPCR. In the two study neighbourhoods, surface water had the highest percentage (96.6%) of *V. cholerae*-positive samples. For open drain and flood water, 70% and 77% of the samples, respectively, were positive for *V. cholerae* (**Table 1**). The sample types with the lowest *V. cholerae* detection were drinking water (5.1%) and street food (5.4%). *V. cholerae* was not detected in any of the shaved ice samples. Chi-squared and the Fisher’s exact test showed that there was no significant difference in the overall percentage of *V. cholerae* positive samples between the two neighbourhoods.

**Table 1:** *V. cholerae* detection by samples type and neighbourhood.

| Percent of Positive Samples (Number of Positive/Number of Samples) |  |  |  |
| --- | --- | --- | --- |
| Sample types | Mukuru Kwa Reuben | Mukuru Kwa Njenga | Total |
| Drinking water<sup>†</sup> | 6.5 (7/107) | 4.0 (6/150) | 5.1 (13/257) |
| Flood water | 60.0 (9/15) | 80.0 (12/15) | 70.0 (21/30) |
| Open drain | 73.3 (33/45) | 80.4 (37/46) | 77.0 (70/91) |
| Public latrine | 20.0 (3/15) | 26.7 (4/15) | 23.3 (7/30) |
| Raw produce | 13.3 (2/15) | 20.0 (3/15) | 16.7 (5/30) |
| Shaved ice | 0 (0/14) | 0 (0/15) | 0 (0/29) |
| Soil | 6.7 (1/15) | 26.7 (4/15) | 16.7 (5/30) |
| Street food* | 7.1 (10/140) | 3.6 (5/137) | 5.4 (15/277) |
| Surface water | 93.3 (14/15) | 100 (14/14) | 96.6 (28/29) |
| Total | 20.7 (79/381) | 20.1 (85/422) | 20.4 (164/803) |

\*Street food includes french fries, githeri (mixture of beans and corn), and mandazi.
†Drinking water includes municipal water, borehole water, water from vendors, and stored drinking water.

### *V. cholerae* concentration in positive quantifiable environmental samples

High concentrations of *V. cholerae* were detected in most of the sample types except drinking water (**Table 2**). For water samples, the highest mean concentration of *V. cholerae* was detected in surface water (3469 gc/mL) and the lowest concentration was observed in drinking water (25.6 gc/mL). The mean concentration of *V. cholerae* in the same sample type varied between the two study neighbourhoods. In Mukuru Kwa Njenga, the mean concentration of *V. cholerae* in surface water was approximately 5-fold higher than the concentration detected in Mukuru Kwa Reuben. Additionally, high concentrations were also detected in drinking water, soil, and Raw produce samples obtained from Mukuru kwa Njenga compared to those from Mukuru Kwa Rueben. Public latrines sampled from Mukuru Kwa Reuben, had a higher mean concentration (6618 gc/swab) of *V. cholerae* compared to Mukuru Kwa Njenga (4811.2 gc/swab). Street food samples collected from Mukuru Kwa Reuben had a slightly higher concentration (587.6 gc/g) than in Mukuru Kwa Njenga (489 gc/g). Due to the small sample sizes of quantifiable results, a two sample t-test did not detect any significant difference in the mean concentrations of *V. cholerae* between two neighbourhoods.

**Table 2.**
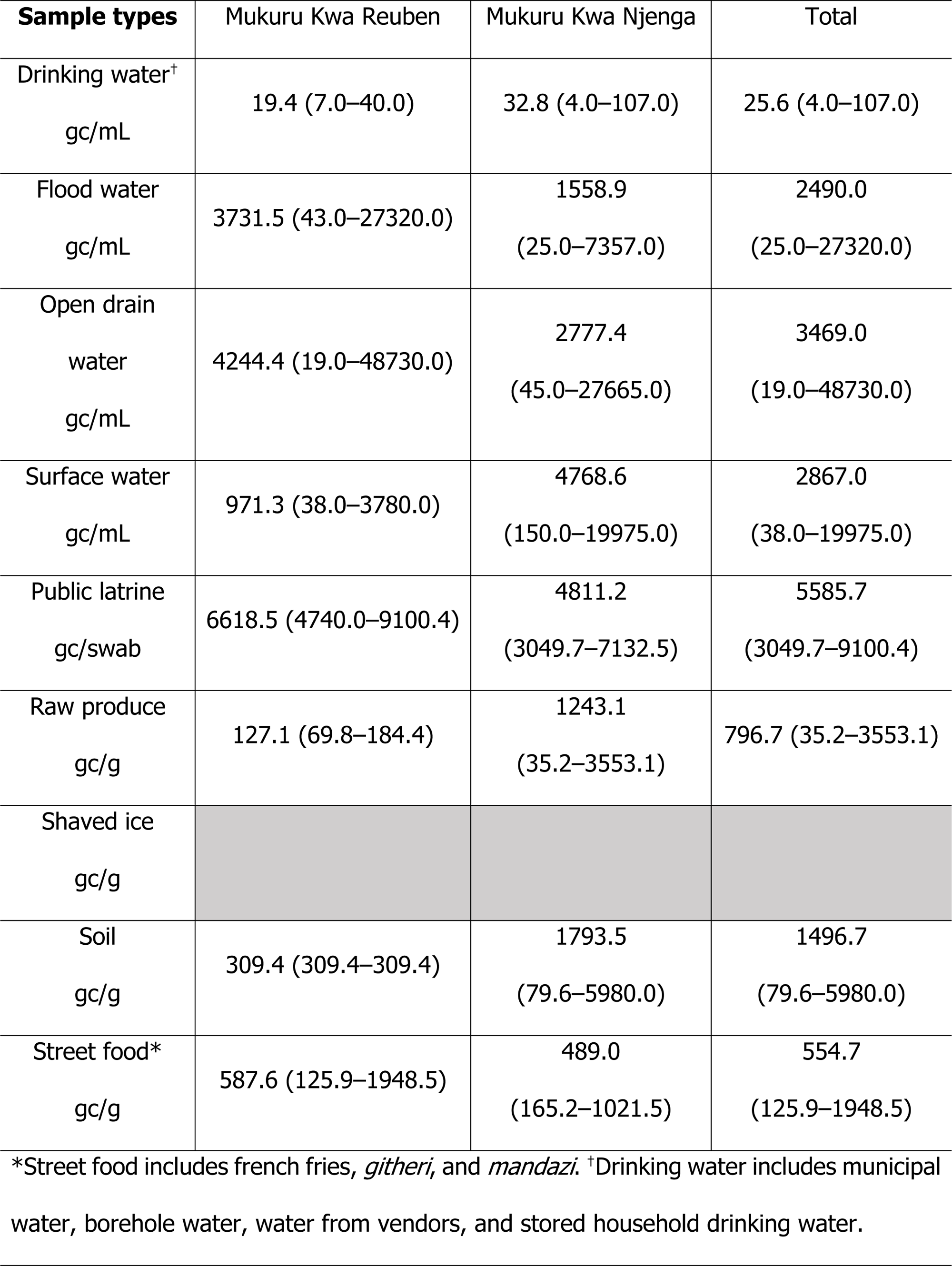
Mean *V. cholerae* concentration in positive quantified samples.

| **Sample types** | Mukuru Kwa Reuben | Mukuru Kwa Njenga | Total |
| --- | --- | --- | --- |
| Drinking water<sup>†</sup><br>gc/mL | 19.4 (7.0–40.0) | 32.8 (4.0–107.0) | 25.6 (4.0–107.0) |
| Flood water<br>gc/mL | 3731.5 (43.0–27320.0) | 1558.9<br>(25.0–7357.0) | 2490.0<br>(25.0–27320.0) |
| Open drain<br>water<br>gc/mL | 4244.4 (19.0–48730.0) | 2777.4<br>(45.0–27665.0) | 3469.0<br>(19.0–48730.0) |
| Surface water<br>gc/mL | 971.3 (38.0–3780.0) | 4768.6<br>(150.0–19975.0) | 2867.0<br>(38.0–19975.0) |
| Public latrine<br>gc/swab | 6618.5 (4740.0–9100.4) | 4811.2<br>(3049.7–7132.5) | 5585.7<br>(3049.7–9100.4) |
| Raw produce<br>gc/g | 127.1 (69.8–184.4) | 1243.1<br>(35.2–3553.1) | 796.7 (35.2–3553.1) |
| Shaved ice<br>gc/g |  |  |  |
| Soil<br>gc/g | 309.4 (309.4–309.4) | 1793.5<br>(79.6–5980.0) | 1496.7<br>(79.6–5980.0) |
| Street food*<br>gc/g | 587.6 (125.9–1948.5) | 489.0<br>(165.2–1021.5) | 554.7<br>(125.9–1948.5) |

\*Street food includes french fries, *githeri*, and *mandazi*. <sup>†</sup>Drinking water includes municipal water, borehole water, water from vendors, and stored household drinking water.

### Temporal trend of *V. cholerae* detection

The detection of *V. cholerae* in different sample types varied between the different time periods of sampling. **Figure 2** shows the temporal distribution of *V. cholerae* positive samples by neighbourhood and for both neighbourhoods combined. In both neighbourhoods, *V. cholerae* detection in surface water and open drain remained high (≥ 50%) throughout the sampling period except in September 2021, when no positive open drain samples were detected. The percentage of positive flood water samples was also high (≥ 80%) throughout the sampling period with the lowest detection (37.5%) observed in August. For drinking water, *V. cholerae* detection was slightly higher during the months of October and November compared to March through April.

**Figure 2:**
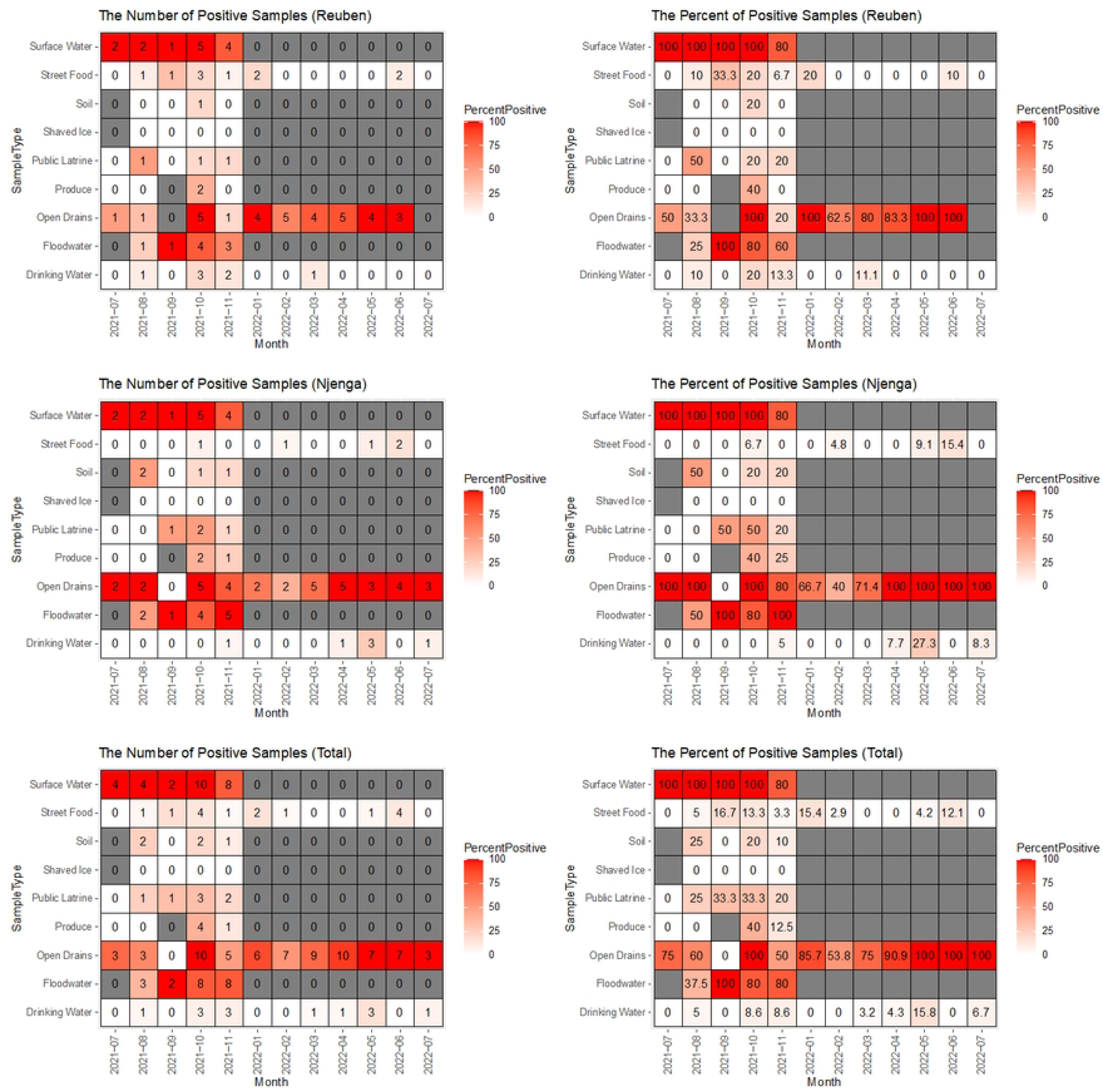
Temporal (monthly) distribution of the number of *V. cholerae* positive samples (left) and percentage of positive samples (right) by neighborhood and sample type, July 2021-July 2022. The color coding indicates represents the percent of positive samples by month in all the plots.

### Environmental pathways with substantial contribution to total exposure to *V. cholerae*

Using the data on *V. cholerae* detection in environmental samples and reported behavior frequency, we estimated exposure to *V. cholerae* through nine environmental pathways. The contribution of a specific pathway to the total exposure to *V. cholerae* was calculated by the exposure to *V. cholerae* from this pathway divided by the sum of the exposures to *V. cholerae* from all pathways. The contribution of the nine pathways, to the total exposure to *V. cholerae*, varied between adults and children.

**Table 3** shows the important pathways that made substantial contributions to the total exposure to *V. cholerae*. **Figure 3** illustrates the contributions of different pathways to the total exposure of *V. cholerae* in both neighbourhoods and between the two age groups. Among children in both neighbourhoods, surface water was the common important pathway of exposure to *V. cholera*e, contributing 84% and 79.7% of the total exposure to *V. cholerae* in Mukuru Kwa Njenga and Mukuru Kwa Reuben, respectively (**Figure 3**). Drinking water also contributed marginally (10.8%) to the total exposure to *V. cholerae* among children in Mukuru Kwa Njenga. Among adults, drinking water had the greatest contribution to total exposure to *V. cholerae*. In Mukuru Kwa Njenga, drinking water had a high (71.1%) contribution to total exposure for adults, while in Mukuru Kwa Reuben four pathways (ingestion of drinking water and street food, contact with surface water, and ingestion of raw produce) substantially contributed to the total exposure (**Figure 3**). The magnitude of the total exposure to *V. cholerae* varied between the two neighbourhoods and among children and adults (represented by the colour in **Figure 3**). The total exposure to *V. cholerae* was higher in Mukuru kwa Njenga compared to Mukuru Kwa Reuben for both adults and children.

**Figure 3.**
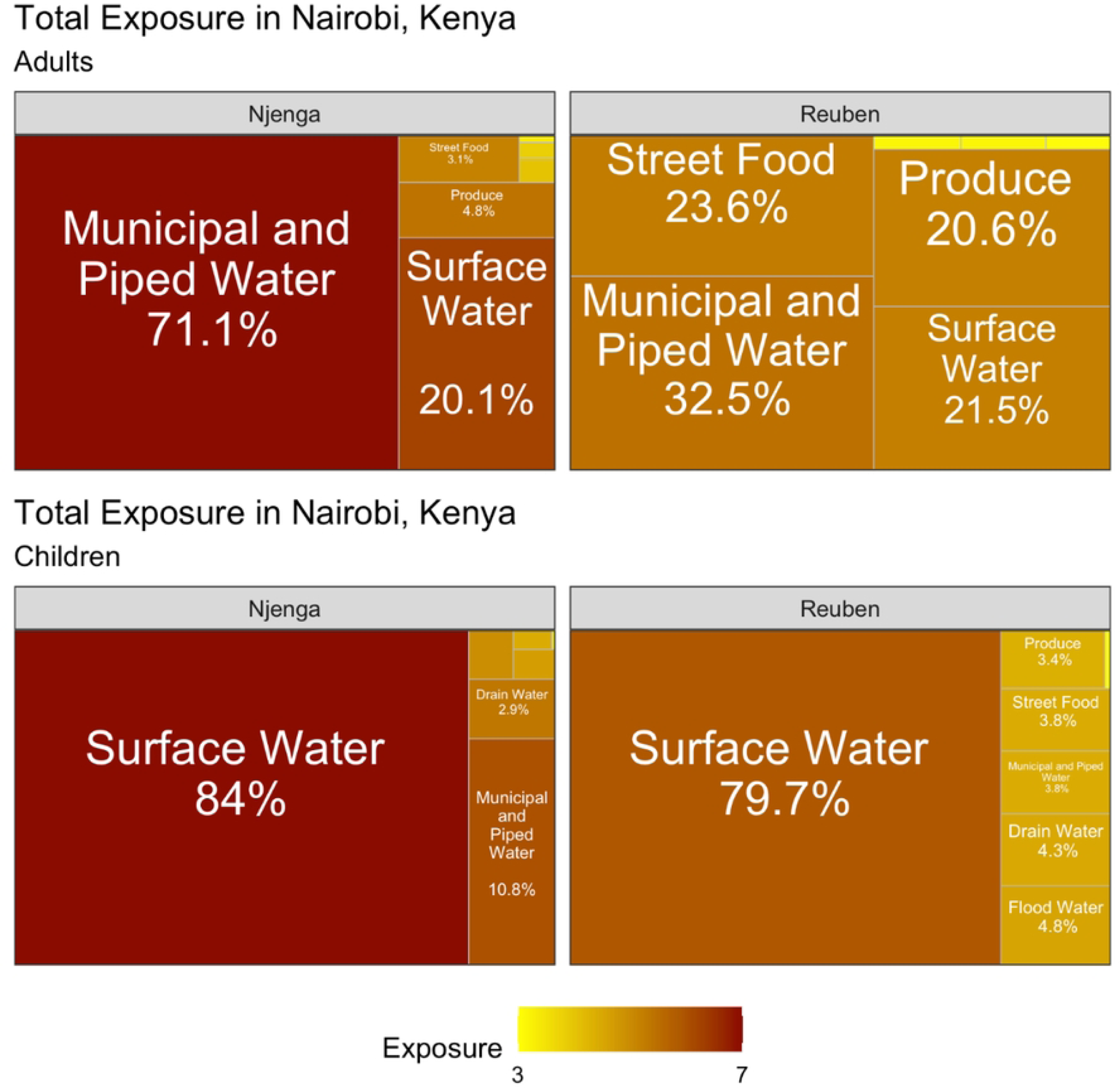
Contributions of different pathways to the total exposure to *V. cholerae* among adults and children in Njenga and Reuben neighbourhoods. The percentage indicates the contribution of each pathway to the total exposure. The color coding represents the magnitude of exposure to *V. cholerae* in every pathway.

**Table 3:**
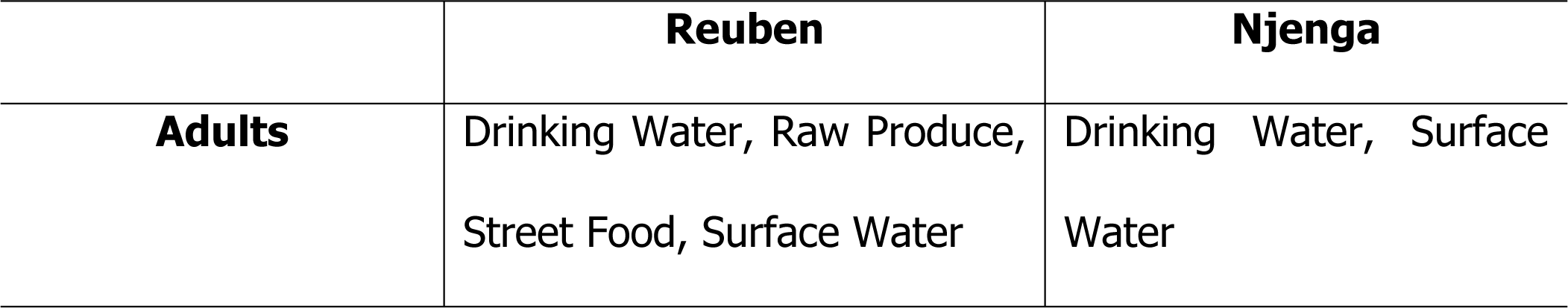
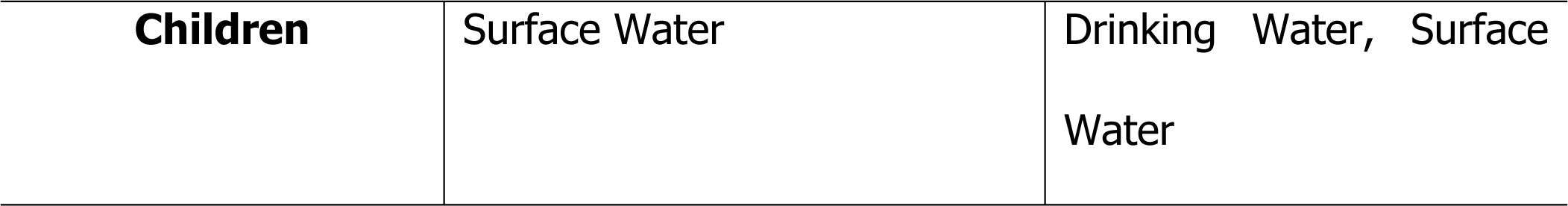
Pathways that made substantial contribution to the total exposure to *V. cholerae* by age group and neighbourhood.

|  | Reuben | Njenga |
| --- | --- | --- |
| Adults | Drinking Water, Raw Produce,<br>Street Food, Surface Water | Drinking Water, Surface<br>Water |
| **Children** | Surface Water | Drinking Water, Surface Water |

## Discussion

Cholera outbreaks continue to be a major health concern in low- and middle-income countries resulting in devastating outcomes, especially among vulnerable populations such as people displaced by natural disasters (e.g., drought and flooding), in refugee camps, and those living in poor informal settlements [30–34]. In Kenya, the most recent cholera outbreak was reported between October 2022 and March 2023 with a CFR of 1.6% [35]. In this study, we aimed to identify the pathways of exposure *to V. cholerae* and the relative contribution of each pathway to the total exposure to *V. cholerae* within the Mukuru informal settlement of Nairobi, which is densely populated and characterized by inadequate sanitation and hygiene.

We identified possible transmission routes and important exposure pathways to *V. cholerae* in the informal settlements of Nairobi. Findings from this study demonstrate the presence of *V. cholerae* in different environmental reservoirs within the informal settlement, which poses a considerable health risk to the populations residing in these areas. Although none of the environmental samples were positive for *V. cholerae* through culture, the hemolysin A gene of *V. cholerae* was detected by qPCR in 20.4% of the samples. The absence of culture-positive samples in this study could imply the presence of viable but non-culturable (VBNC) or conditionally viable environmental cells (CVEC) of *V. cholerae* [36]. A study conducted in Uganda also reported that none of the surface water samples analysed for *V. cholerae* were culturable since all the samples were *Crystal VC* dipstick negative [37]. These findings demonstrate the importance of utilizing different detection methods because dependence on culture-based methods alone may be unreliable. In the VBNC form, *V. cholerae* reduces metabolic activity due to environmental stress and nutrient deprivation and is not detectable through culture-based techniques [38] but remains viable for more than a year [39]. The detection of *V. cholerae* through qPCR in this study could be an indication of the widespread presence of VBNC in this informal settlement and underscores the need for utilizing molecular methods for the detection of *V. cholerae*, especially in cholera endemic areas. The presence of *V. cholerae* virulence genes in environmental reservoirs in Mukuru Informal settlement is of grave concern considering that VBNC *V. cholerae* cells can become infectious after ingestion by humans [40] and/or in favourable conditions [41,42], which could result in cholera outbreaks.

In the current study, *V. cholerae* was detected in > 96% of surface water samples, which was much higher than the 10.8% detection rate reported in Uganda [37]. In Dhaka, Bangladesh, *V. cholerae* was detected in a large percentage of open drain samples (> 93%) [29], which is comparable to what was observed in this study. The high concentrations of *V. cholerae* in environmental water reservoirs, including surface water, open drain water, and flood water, observed in this study highlight the poor sanitation conditions in the informal settlements. The surface water in these areas is part of the Ngong River [43], and considering that some households are built on the River banks [44], there is a possibility that fecal waste from the households ends up in open drains and into the river. The detection of *V. cholerae* in the environmental samples throughout the study period may be due to the continuous poor disposal of fecal matter into the open drains and surface waters and the long-term persistence of *V. cholerae* in the environmental reservoirs.

Given that cholera outbreaks have previously been associated with contaminated drinking water [43,45,46], the presence of *V. cholerae* in drinking water and street food is worrying since it can lead to possible large outbreaks in communities that frequently buy street foods and rarely treat drinking water at home.

Although there was a difference in the contribution of different pathways to the total exposure to *V. cholerae* among adults, four pathways (drinking water, surface water, street food and produce) were prominent. Drinking water made a substantial contribution to the total exposure for adults, especially in Mukuru Kwa Njenga. While the concentration of *V. cholerae* in drinking water was low compared to environmental waters, the high contribution to *V. cholerae* exposure could be attributed to the frequent direct consumption of drinking water. The exposure to *V. cholerae* among adults through drinking water is an indication of consumption of water that is not treated or inadequately treated, which is common in the informal settlement [24].

For children in both neighbourhoods, surface water had the highest contribution to total exposure to *V. cholerae*. In addition to the high concentration of *V. cholerae* in surface water, children may be exposed to *V. cholerae* when playing around or crossing the surface water (Ngong River) through informally constructed bridges. This underscores the need for better infrastructure in the area and sensitization of residents on the impact of proper waste management. Our findings indicate that levels of absolute exposure to *V. cholerae* were higher in Mukuru Kwa Njenga for both children and adults. This could be caused by a higher concentration of *V. cholerae* in the environment and more frequent contact with the environment in Mukuru kwa Njenga. This could be attributed to the high population in Mukuru Kwa Njenga and the fact that the WASH infrastructure is much better in Mukuru Kwa Reuben. These findings show that multiple pathways contribute to exposure to *V. cholerae* among vulnerable populations, therefore, oral cholera vaccination and improvement of WASH infrastructure and services are critical for the prevention and control of cholera outbreaks.

Our findings also showed that in the three sample types (drinking water, open drains, and street foods) collected for one year, *V. cholerae* detection peaked during two time periods; May and September–October, which are part of the rainy/wet seasons in Kenya. Similar findings were observed in Uganda, where the detection of *V. cholerae* peaked in the months of March–July, which corresponds to wet seasons [37]. *V. cholerae* contamination in the environment may be more likely to be transferred to street food because of poor hygiene and food handling practices during months when there is more *V. cholerae* in the environment.

Although *V. cholerae* was detected in drinking, street food and produce no cholera outbreak was reported during the study period. This could be because the contamination levels were not sufficiently high to cause infection in most exposure incidents and there may be under-ascertainment and underreporting of cholera in these poor neighbourhoods. However, in October 2022, two months after the completion of the study, there was a cholera outbreak with a high burden of cases reported in low-resource settings, including the study sites. The absence of reported cholera cases during the study period could be an indication that traditional case-based passive surveillance may not be sufficient to indicate the true circulation of *V. cholera* in the population. Environmental surveillance may be useful in these settings to complement conventional surveillance to monitor endemic and seasonal epidemic cholera. This is critical considering studies have reported that major cholera outbreaks are preceded by an increase in the presence of *V. cholerae* in the environment [47].

### Strengths and Limitations

This study identified the probable routes of transmission of *V. cholerae* in an informal settlement and quantitatively assessed exposure to *V. cholerae* through different environmental pathways for children and adults. Three sample types (drinking water, street food, and open drain water) samples were collected weekly over a period of 12 months to examine temporal patterns in *V. cholerae* detection. This study was conducted in several zones within cholera-prone informal settlements and provide insights about spatial variation in the presence of *V. cholerae* in environmental reservoirs across small geographic areas. We also utilized both molecular and conventional methods in the detection of *V. cholerae* in environmental samples.

This study has several limitations. First, there were only five sampling points for every sample type per neighbourhood, which may not have been sufficient to represent the entire neighbourhood. Second, the two study neighbourhoods form a small section of a larger informal settlement and the findings may not be generalizable to other parts of the informal settlements. Third, whereas we measured exposure, it does not directly translate to infection or disease risk.

## Conclusion

This study identified multiple environmental pathways of exposure to *V. cholerae* in two informal settlements in Nairobi, Kenya. Because more than one pathway substantially contributed to the total exposure to *V. cholerae* in these neighbourhoods, these findings triggered an oral cholera vaccine campaign and distribution of safe water storage containers and point of use drinking water chlorination kits (aquatabs) as immediate prevention and control measures. The study findings also demonstrate the importance of proper and sustainable water, sanitation and hygiene infrastructure and services in high-risk areas. Seasonal variation in the detection of *V. cholerae* was observed. Our study findings also demonstrate the potential use of environmental surveillance to monitor temporal trends in cholera incidence and provide early warnings of cholera outbreaks. There is also a need to conduct long-term surveillance to determine the impact of climate change on the presence or persistence of *V. cholerae* in the environment. Further studies should also be undertaken to understand the phylogenetic relatedness and antimicrobial resistance in *V. cholerae* from both clinical and environmental sources within the informal settlement, and their role in sporadic cholera outbreaks in the settlement.

## Data Availability

All data has been provided as part of the submitted article.

## Acknowledgement

We would like to thank the community health volunteers who helped with conducting the transect walks and the fieldworkers who were involved in sample collection and processing. We also want to thank the Director General of Kenya Medical Research Institute for the support in undertaking the study.

## Author contributions

**Conceptualization**: Kelvin Kering, Cecilia Mbae, Habib Yakubu, Pengbo Liu, Wondwossen Gebreyes, Christine Moe, and Samuel Kariuki.

**Data curation**: Kelvin Kering, Michael Mugo, Beatrice Ongadi, Peter Muturi, Georgina Odityo and Sarah Durry

**Methodology**: Kelvin Kering, Habib Yakubu, Aniruddha Deshpande, Pengbo Liu and Sarah Durry

**Data analysis**: Kelvin Kering, Yuke Wang, Pengbo Liu and Aniruddha Deshpande.

**Supervision**: Cecilia Mbae, Habib Yakubu, Pengbo Liu, Christine Moe and Sarah Durry.

**Project administration**: Cecilia Mbae

**Funding acquisition**: Wondwossen Gebreyes, Christine Moe and Samuel Kariuki.

**Writing-original draft**: Kelvin Kering, Yuke Wang, and Samuel Kariuki

**Writing-review and editing**: Kelvin Kering, Cecilia Mbae, Michael Mugo, Beatrice Ongadi, Georgina Odityo, Peter Muturi, Yuke Wang, Habib Yakubu, Pengbo Liu, Sarah Durry, Aniruddha Deshpande, Wondwossen Gebreyes, Christine Moe, and Samuel Kariuki

